# Linking Genotype to Phenotype: Further Exploration of Mutations in SARS-CoV-2 Associated with Mild or Severe Outcomes

**DOI:** 10.1101/2022.04.15.22273922

**Authors:** Roshna Agarwal, Tyler Leblond, Erin M McAuley, Ezekiel J Maier, Martin Skarzynski, Jameson D Voss, Shanmuga Sozhamannan

## Abstract

We previously interrogated the relationship between SARS-CoV-2 genetic mutations and associated patient outcomes using publicly available data downloaded from GISAID in October 2020 [1]. Using high-level patient data included in some GISAID submissions, we were able to aggregate patient status values and differentiate between severe and mild COVID-19 outcomes. In our previous publication, we utilized a logistic regression model with an L1 penalty (Lasso regularization) and found several statistically significant associations between genetic mutations and COVID-19 severity. In this work, we explore the applicability of our October 2020 findings to a more current phase of the COVID-19 pandemic.

Here we first test our previous models on newer GISAID data downloaded in October 2021 to evaluate the classification ability of each model on expanded datasets. The October 2021 dataset (n=53,787 samples) is approximately 15 times larger than our October 2020 dataset (n=3,637 samples). We show limitations in using a supervised learning approach and a need for expansion of the feature sets based on progression of the COVID-19 pandemic, such as vaccination status. We then re-train on the newer GISAID data and compare the performance of our two logistic regression models. Based on accuracy and Area Under the Curve (AUC) metrics, we find that the AUC of the re-trained October 2021 model is modestly decreased as compared to the October 2020 model. These results are consistent with the increased emergence of multiple mutations, each with a potentially smaller impact on COVID-19 patient outcomes. Bioinformatics scripts used in this study are available at https://github.com/JPEO-CBRND/opendata-variant-analysis. As described in Voss et al. 2021, machine learning scripts are available at https://github.com/Digital-Biobank/covid_variant_severity.

## Introduction

The Global Initiative on Sharing Avian Influenza Data (GISAID) is a popular and publicly available repository that houses SARS-CoV-2 sequencing data from the global community. GISAID stores SARS-CoV-2 genomic data and sequencing metadata—including sequencing technology and assembly method—as well as some patient metadata, including high-level patient outcomes, region of origin, age, gender and date of collection.

We have previously used the high-level patient metadata submitted by GISAID users to differentiate between severe and mild patient outcomes. Briefly, we aggregated patient outcomes into “Mild” outcomes (e.g., Outpatient, Asymptomatic, Mild, Home/Isolated/Quarantined, Not Hospitalized) or “Severe” outcomes (e.g., Hospitalized (including severe, moderate, and stable) and Deceased (Death)). We excluded entries whereby the severity of the condition could not readily be discerned, (e.g., retesting, screened for travel, not vaccinated, moderate covid, and live). Following this approach, in Voss et al., 2021, of 3,637 SARS-CoV-2 samples from GISAID with patient outcome metadata, 2,870 were classified as severe patient outcomes and 767 as mild.

Using this patient outcome classification described previously (Voss et al., 2021), we showed logistic regression models that included viral genomic mutations outperformed other models that used only patient age, gender, sample region, and viral clade as features. Among individual mutations, we found 16 SARS-CoV-2 mutations that have ≥ 2-fold odds of being associated with more severe outcomes, and 68 mutations associated with mild outcomes (odds ratio ≤ 0.5). While most assessed SARS-CoV-2 mutations are rare, 85% of genomes had at least one mutation associated with patient outcomes.

## Methods

Metadata preprocessing, cohort building, mutation and metadata modeling, data visualization, and statistical analyses were all done according to procedures in Voss et al., 2021 and are diagramed in Figure 1. Briefly, an export of raw GISAID SARS-CoV-2 data was curated using Nexstrain’s ncov-ingest shell scripts [2] and FASTA sequences were parsed from the data export using Python (version 3.8.10). Of the 4,646,285 samples available in GISAID through ncov-ingest on October 26^th^ 2021, we used a subset of 53,787 samples with patient outcome metadata for our analyses. We utilized a total of 29,359 severe and 24,428 mild samples for our analyses. FASTA sequences were aligned to the reference sequence, Wuhan-Hu-1 (NCBI: NC_045512.2; GISAID: EPI_ISL_402125) using Minimap2. Resulting VCF (Variant Call Format) files were merged using bcftools and annotated using SnpEff and filtered using SnpSift. We applied a similar bioinformatics analysis pipeline to Rayko et al [7] and is available under the scripts directory at https://github.com/JPEO-CBRND/opendata-variant-analysis.

**Figure 1.**
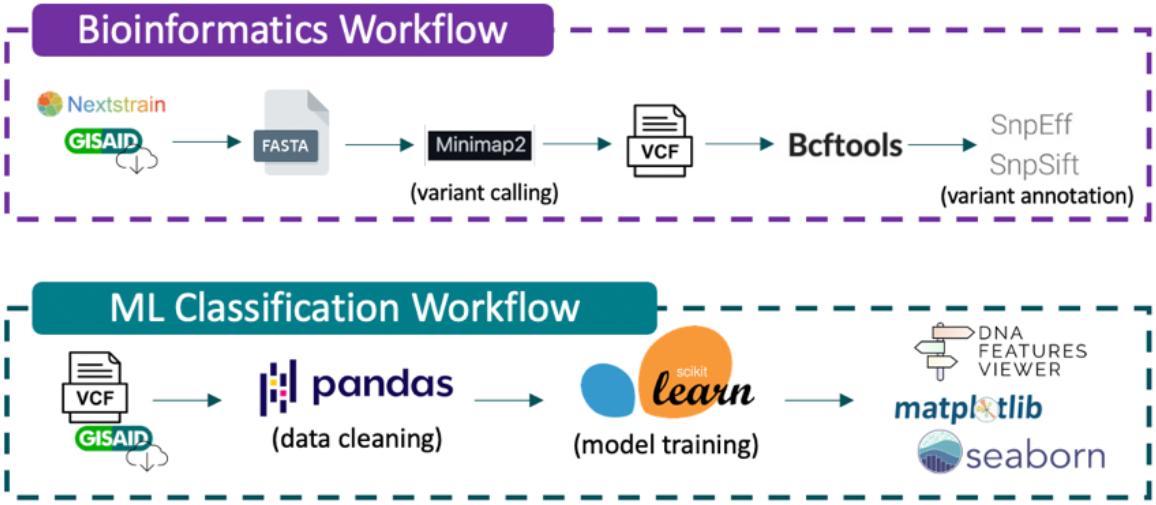
Bioinformatics and Machine Learning Classification workflows used for SARS-CoV-2 genetic variant calling and annotation, and mutation and metadata modeling based on procedures used in Voss et al., 2021.

The machine learning classification workflow, available at https://github.com/Digital-Biobank/covid_variant_severity, first runs 00_long.py to join and converts annotated VCF files to a single parquet file. Next, 00_red.py is run against GISAID metadata and aggregates patient outcomes into positive (‘Mild’), or negative (‘Severe’) outcomes and stores the classification into a recode dictionary. 01_wide.py, 02_var-freq.py, 02_join.py, and 03_clean.py are consecutively run for pivoting to wide format, deriving mutation frequencies per sample, joining the VCF parquet file with GISAID patient data, and further parsing. 04_logit.py runs training and testing for the logistic regression models. Finally, 05_plot-variants.py is run to derive a table of highest and lowest odds of mutations being classified into mild or severe outcomes.

Scikit-learn [4] was used to fit logistic regression models with the L1 penalty (Lasso regularization) and the default regularization strength (C = 1) to the patient (rows) and mutation (columns). A train/test split was created on the data (67% train, 33% test). The training data were split into five cross-validation folds using the Scikit-learn stratified K-fold cross-validation generator. Test data was only used for evaluating the performance of trained models. Models were persisted as pickle files using joblib. Scatter and bar plots were created using Pandas [3], Matplotlib [5] and Seaborn [6]. ROC curves were plotted using Scikit-learn [4], and Matplotlib [5]. The versions of all tools used for bioinformatics and machine learning analysis are shown in Table 1.

**Table 1.**
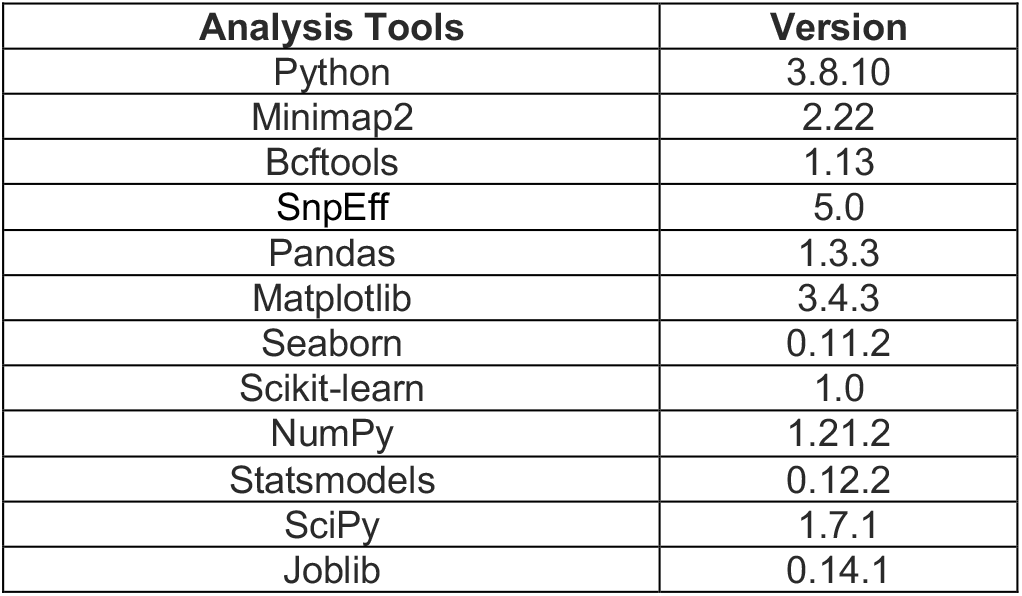
Versioning for Python utilities and third-party software tools used in shell scripts for machine learning classification and bioinformatics workflows.

Similar to previous work (Voss et al., 2021), we trained a total of five logistic regression models using different input features. For each model, Scikit-learn [4] was used to calculate the area under the curve (AUC), a measure of goodness of fit of a binary classification model. AUC confidence intervals, P-values, and diagnostic odds ratios (OR) were calculated using NumPy [9] for each of the five logistic regression models. The Scikit-learn implementation of logistic regression does not provide ORs or P-values for individual variables. ORs and Chi-square test P-values for the association of mutations with Severe and Mild outcomes (Figure 5) were calculated from mutation count data using Statsmodels and SciPy respectively [8]. Mutation frequency was calculated using Pandas [3].

## Results

### I. Validation of Previous Methods

Before testing and retraining the logistic regression models on newer data downloaded in October 2021, we first reproduced the previous results using the same dataset (Voss et al., 2021). Reassessing the previous results, the model using age, gender, region, and variants as features had the highest AUC (0.91) and accuracy (91%), followed by models that use fewer features (age/gender/region/clade, age/gender/region, age/gender, and age). The SARS-CoV-2 mutations significantly associated with disease severity identified previously, were also replicated in this reanalysis (Voss et al., 2021).

### II. Evaluation of Model Performance on the Expanded GISAID Dataset

Next, we evaluated the classification performance of the previous logistic regression models (Voss et al., 2021) on the newer, expanded October 26^th^, 2021 GISAID dataset. For this evaluation, the genetic mutations included in the expanded October 2021 dataset was limited to match the feature space of the trained previous logistic regression models (Voss et al., 2021). Therefore, mutations observed in the October 2021 dataset, but not the October 2020 dataset, were not included in this test dataset. The performance of the trained Voss et al., 2021 logistic regression models was evaluated on the test split (67% train, 33% test) of the expanded October 2021 dataset.

Figures 2 and 3 below show comparisons of ROC curves and model performance statistics for the Voss et al., 2021 logistic regression models on the original October 2020 dataset and the expanded October 2021 dataset. An overall decline in performance is observed for the previous models when applied to the expanded October 2021 dataset. Notably, testing the original models on the October 2021 dataset reveals a decrease in performance for models that include the region feature (AGRV AUC: 0.911 vs. 0.580; AGRC AUC: 0.818 vs. 0.571; AGR AUC: 0.817 vs. 0.564).

**Figure 2.**
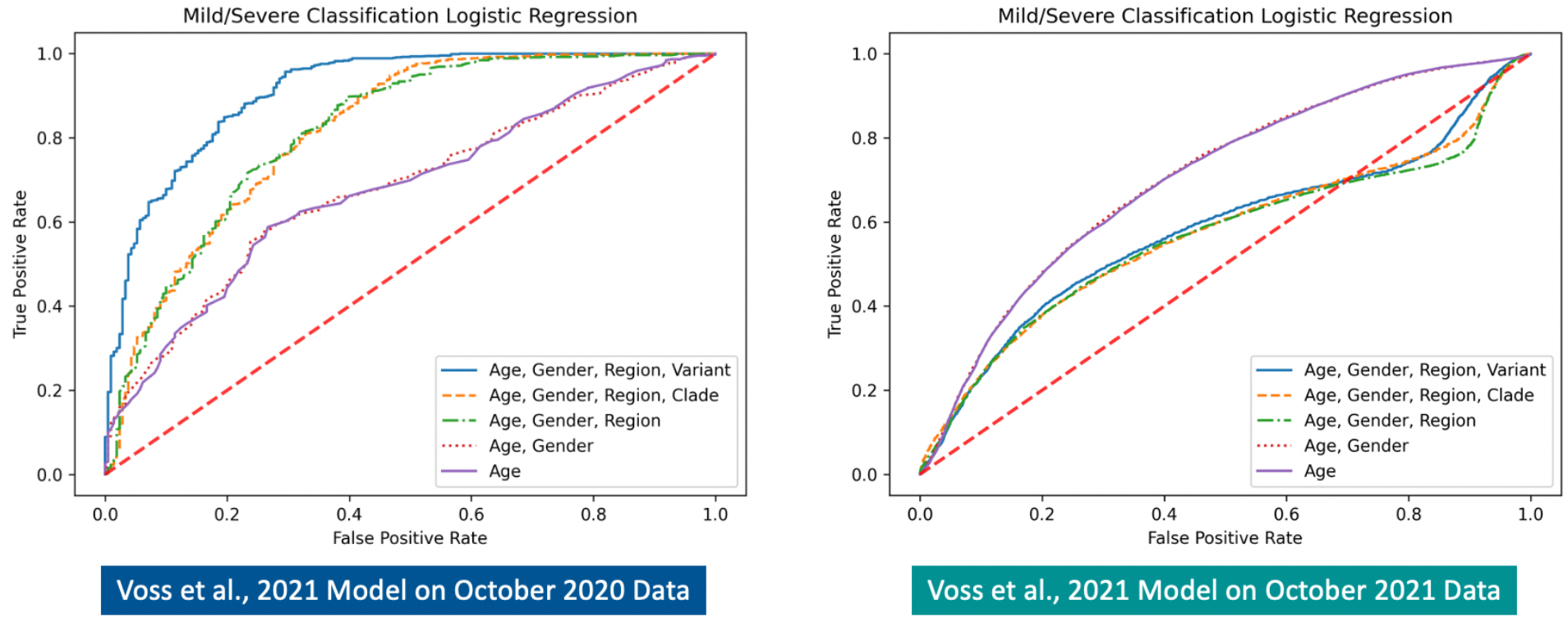
Comparison of ROC curves for the previous (Voss et al.,2021) logistic regression models tested on the October 2020 dataset (left) and the expanded October 2021 dataset (right). A decrease in model performance is observed on the expanded October 2021 dataset.

**Figure 3.**
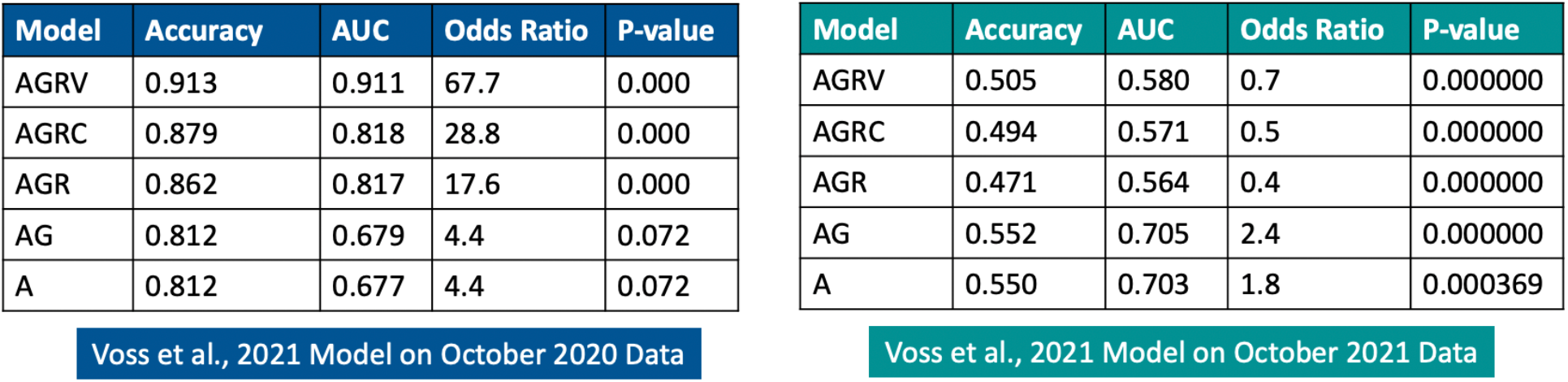
Comparison of previous (Voss et al.,2021) model performance statistics (left) and the previous models run on the expanded October 2021 dataset (right)

### III. Re-Analysis on Expanded GISAID Dataset

To further investigate our previous findings [1], the nested logistic regression models were retrained on the larger October 26^th^, 2021 GISAID SARS-CoV-2 dataset. Model retraining was done using the train split of the expanded October 2021 dataset, while performance was evaluated using the test split (67% train, 33% test). The performance of the retrained models was then compared to the logistic regression models trained on the October 20^th^, 2020 GISAID SARS-CoV-2 dataset. ROC curves for the retrained models are shown in Figure 4A (left). The model using age, gender, region, and variants (AGRV) as features continues to show the best performance, as observed previously (Voss et al, 2021). The retrained AGRV model metrics, shown in Figure 4B (right), have an overall decrease in model accuracy (from 0.913 to 0.810) and AUC (from 0.911 to 0.885) as compared to previously published data (Voss et al., 2021) shown in Figure 3A (left). This decrease in retrained AGRV model performance may indicate a modest reduction in power to distinguish between severe and mild outcomes in the expanded October 2021 dataset, or may be explained by inconsistent case severity definitions between the 2020 and 2021 datasets. SARS-CoV-2 mutations most associated with severe and mild outcomes, as measured by odds ratio, from the previous study (Voss et al., 2021) and the expanded October 26^th^, 2021 GISAID dataset are shown in Figure 5. None of the mutations with the highest (n=20) and lowest (n=20) odds of being associated with severe or mild outcomes from earlier findings (Voss et al., 2021) are also identified in the top 40 mutations of the larger October 2021 dataset analysis. While there is no overlap in the mutations with the highest and lowest mutations between earlier and the expanded October 26^th^, 2021 GISAID dataset, 10 of the top 20 mutations most associated with severe outcomes from the earlier study (Voss et al., 2021) are also significantly associated with severe outcomes (OR ≥ 2, P-value ≤ 0.05) in the expanded October 2021 dataset. Similarly, 14 of the top 20 mutations most associated with mild outcomes (OR ≤ 0.5, P-value ≤ 0.05) from Voss et al., 2021 are also significantly associated with mild outcomes in the expanded October 2021 dataset.

**Figure 4.**
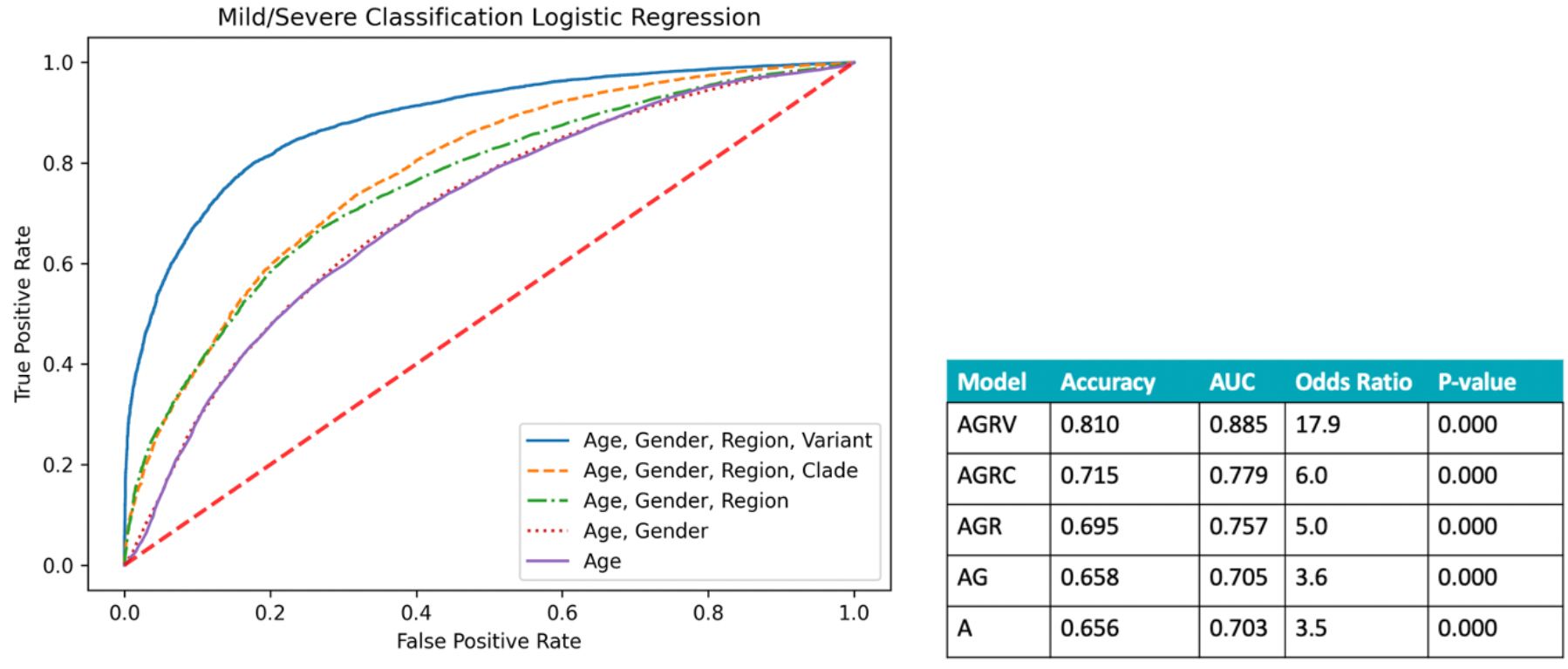
ROC curves after retraining on the expanded October 26^th^, 2021 GISAID dataset (left) and updated model performance metrics (right). Previous (Voss et al., 2021) and retrained model performance is superior when genomic mutations are included as features. A general decrease in model performance was observed for the retrained models on the October 2021 dataset in comparison to the previous models, which used the October 20^th^, 2020 dataset.

**Figure 5.**
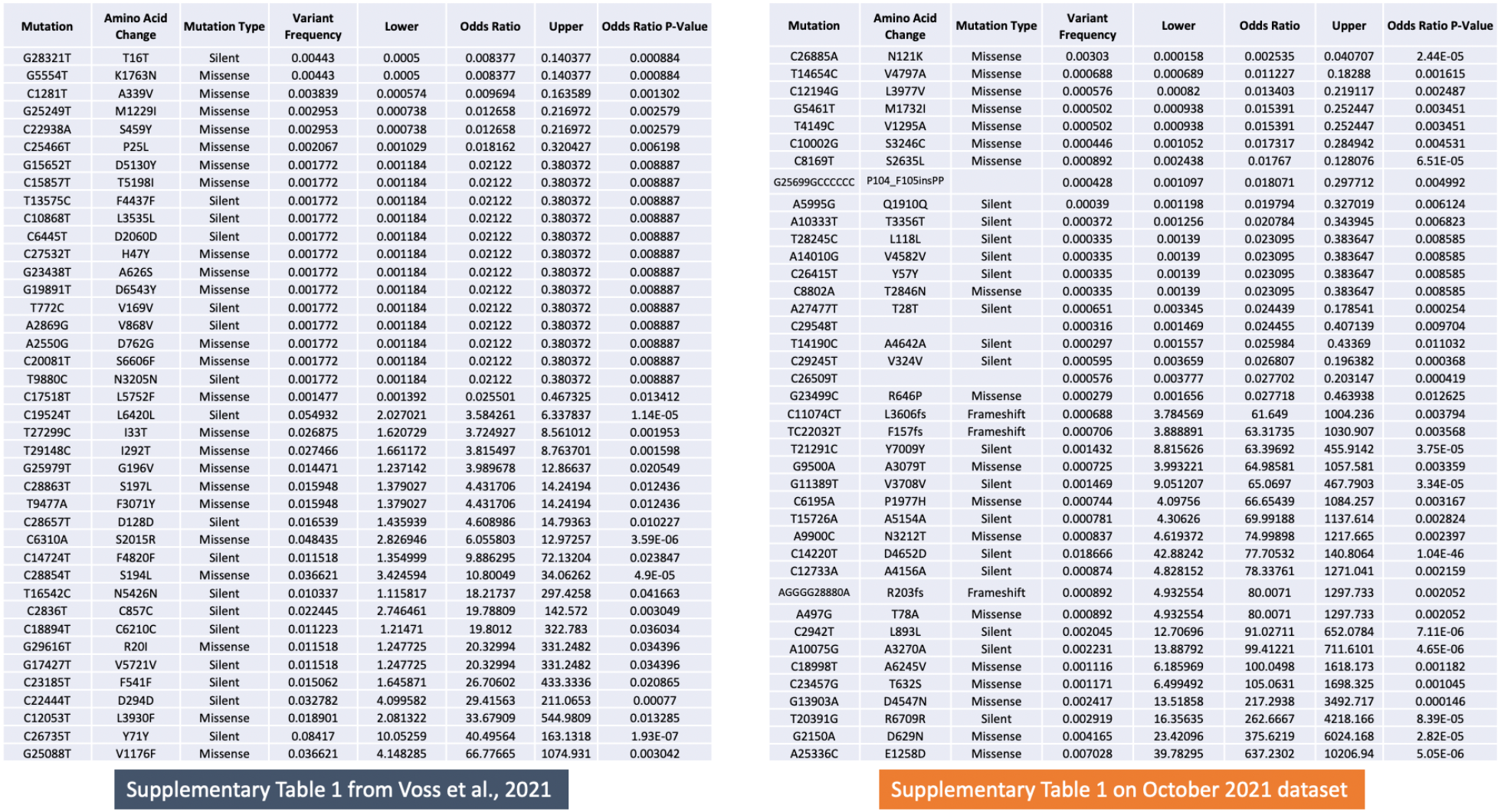
The 40 SARS-CoV-2 mutations with the highest (n=20) and lowest (n=20) odds of being associated with severe or mild outcomes from the previous study (Voss et al., 2021) on the left, and this updated analysis using a GISAID dataset from October 2021 (right). Mutations are ordered by odds ratio, and amino acid change, variant frequency, confidence intervals, and P-values are provided.

### IV. Expansion of Analytical Models on Original Dataset

We explored additional machine learning binary classifiers, including Random Forest, Naïve Bayes, and Neural Network algorithms, and compared their performance to the logistic regression model. Similar to our previous study (Voss et al., 2021), 3,386 samples were used for this analysis, with 2,694 associated with severe outcomes and 692 with mild patient outcomes. Age, gender, region, and variants (AGRV) were used as features for each model. A stratified 67% train and 33% test data split was created using Sci-kit learn model selection module [4], and a 5-fold cross-validation was performed to select the best parameters for each model. Random Forest, Naïve Bayes, and Neural Network algorithms were run using Sci-kit learn ensemble, naïve_bayes, and neural_network modules respectively. As shown in Figure 6, the random forest model outperformed the other models, including the logistic regression model, with an AUC of 0.936, accuracy of 0.918 and odds ratio of 116.7.

**Figure 6.**
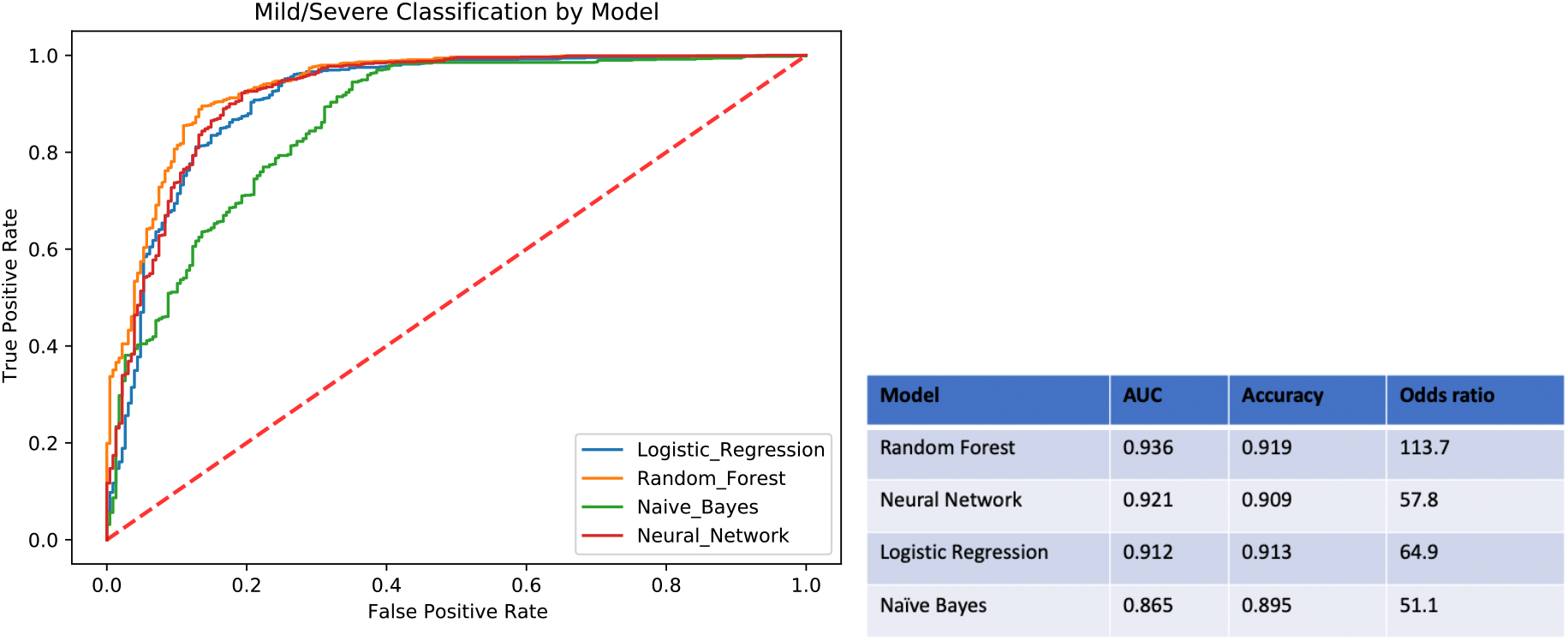
Comparison of alternative machine learning algorithms for binary classification (Random Forest, Neural Network, and Naïve Bayes) to Logistic Regression using October 20^th^, 2020 dataset and AGRV feature set (age/gender/region/variant). The random forest model had the highest performance and the naïve bayes model had the lowest performance.

## Discussion

Herein we further investigated the results published earlier (Voss et al., 2021) by evaluating the performance of the logistic model for classifying COVID-19 severity on a larger SARS-CoV-2 dataset curated from GISAID on October 26^th^, 2021. Testing the previous (Voss et al. 2021) and retrained logistic regression models on the expanded October 2021 GISAID dataset revealed a general reduction in model performance. Previous results (Voss et al., 2021) indicated that using age, gender, region, and variant (AGRV) features enabled the best performance for COVID-19 patient outcome classification. The performance of the retrained logistic regression models on the October 2021 GISAID dataset continues to demonstrate that inclusion of genomic mutation features improves classification of COVID-19 patient outcomes. A modest 2.7% reduction in AUC was observed for the AGRV logistic regression models re-trained on the October 2021 dataset in comparison to the AGRV logistic regression model from the previous study.

The performance of the previously (Voss et al. 2021) trained AGR (age/gender/region), AGRC (age/gender/region/clade), and AGRV (age/gender/region/variant) models was severely degraded when tested on the expanded October 2021 dataset. This diminished performance could be the result of new individual mutations and SARS-CoV-2 variants that weren’t observed in the October 2020 dataset, and vaccination and treatment advances from October 2020 to October 2021. Notably, the AGR, AGRC, and AGRV models all included region in their feature set, and the differential availability of vaccination and treatment advances would be expected to impact region features. We further investigated the potential deleterious impact of the region feature on the previous (Voss et al. 2021) models when applied to the expanded October 2021 dataset by testing an AGV (age/gender/variant) model and comparing its performance to the AG and AGRV models. We find that the AGV model has an AUC and accuracy that is similar to the AG model, rather than the AGRV model (AUC: AG=0.705, AGV=0.709, AGRV=0.580; Accuracy: AG=0.552, AGV=0.550, AGRV=0.505).

Figure 5 lists the top twenty mutations with highest and lowest odds ratios for association with severe and mild outcomes from the October 2020 and expanded October 2021 datasets. The top three mutations most associated with severe outcomes in the October 2021 dataset are T20391G (ORF1ab: R6709R), G2150A (ORF1ab: D629N), and A25336C (S: E1258D). E1258D, a missense mutation located at the cytoplasmic tail of the spike protein [10] that is observed with a frequency of 0.7%, has the highest odds (OR=637.23) for association with severe outcomes. Interestingly, an independent study using machine learning approaches to model COVID-19 disease outcomes also identified E1258D as a key predictor of disease severity [11]. This linage independent mutation is recurrent and arises independently in samples taken from donors and cell lines, indicating potential selection in host environments [12]. The three mutations most associated with mild outcomes include C26885A (M: N121K; OR=0.0030), T14654C (ORF1ab: V4797A), and C12194G (ORF1ab: L3977V). N121K, a missense mutation in the membrane protein observed with frequency of 0.3%, is most associated (OR = 0.0025) with mild outcomes. The presence of this mutation was identified as a key predictor of asymptomatic outcomes in previous machine learning modeling of COVID-19 disease outcomes [13].

In a comparison of machine learning algorithms, we found Random Forest to be the best performing algorithm for classification. The superior performance of the random forest model over the logistic regression and naïve bayes models may indicate the presence of nonlinear interactions between features (e.g., SARS-CoV-2 mutations). Indeed, Random Forest is a well utilized machine learning method for genomic data analysis because this algorithm applies well to problems with many more features than observations and accounts for interactions between features [14][15]. In addition to exploring machine learning algorithm options, the space of outcome variables can also be explored. For example, a promising direction for future modeling is the investigation of mutations associated with transmissibility.

The utilization of supervised learning machine learning poses a limitation in our analysis. Since labeled outcomes are required to train these models, the number of samples available for training is reduced by 99% (53,787 of 4,646,285). In addition, machine learning models trained on older samples may not be sufficiently exposed to new mutations. For example, while many of the more than 50 mutations present in Omicron were observed previously in other variants of concern, some Omicron mutations were rare or previously unobserved and many previously observed mutations hadn’t co-occurred in the same samples [16]. Supervised machine learning models cannot effectively utilize previously unobserved mutations and mutations combinations because parameters have not been fit for these features. A promising approach for addressing these limitations is semi-supervised learning. This machine learning approach uses both labeled data and unlabeled data for model training. Semi-supervised learning may outperform supervised learning approaches when the amount of unlabeled data is much larger than labeled data [17]. Within the field of genomics, recent example uses of semi-supervised learning include microRNA classification [18], somatic genomic variant classification [19], and identify disease associated genes [20].

## Data Availability

All data produced in the present study are available from GISAID upon request to the data owners and in accordance with appropriate data use agreements.

https://www.gisaid.org/

## Disclaimer

*The views and opinions in this article are those of the author and do not necessarily reflect that of the JPEO-CBRND, the U*.*S. Army, the Department of Defense, or the U*.*S. Government. Clearance DSS 22055*.

